# Refining the Prothrombotic State and Prognosis in Atrial Fibrillation With Left Atrial Appendage 3D Echocardiography

**DOI:** 10.1101/2024.01.09.24301079

**Authors:** Laurie Soulat-Dufour, Sylvie Lang, Théo Simon, Stephane Ederhy, Saroumadi Adavane-Scheuble, Marion Chauvet Droit, Elodie Capderou, Camille Arnaud, Eleonore Sotto, Raphael Cohen, Thibault d’Izarny Gargas, Aliocha Scheuble, Nadjib Hammoudi, Anne-Sophie Beraud, Karima Addetia, Franck Boccara, Roberto M. Lang, Ariel Cohen

## Abstract

**BACKGROUND:** Left atrial (LA) volume is an echocardiographic marker of remodeling, thromboembolic risk, and prognosis in atrial fibrillation (AF); limited data are available on LA appendage (LAA) characterization beyond morphology. We sought to evaluate LAA characteristics in 2-dimensional (2D) and 3-dimensional (3D) transesophageal echocardiography (TEE) and the correlation with LA/LAA prothrombotic state and prognosis.

**METHODS:** We prospectively studied 206 hospitalized patients with AF using 2D transthoracic echocardiography (TTE) and 2D/3D TEE of the LAA ≤24 hours from admission. Patients were divided according to the presence or absence of LAA sludge and/or thrombus. Data on clinical events were collected for 2 years.

**RESULTS:** Patients with LAA sludge/thrombus (n=35) on admission had higher LA volumes, lower left ventricular ejection fraction, lower LAA emptying and filling flow velocity, larger 2D LAA measurements (2D LAA ostium diameter, 2D LAA area) as well as larger 3D LAA measurements (higher 3D LAA volumes (LAAV), higher 3D end-systolic [ES] LAA ostium area), and more frequently non-chicken wing morphology. On multivariable logistic regression analysis, LAA filling flow velocity and 3D ES LAAV were associated with the presence of LAA sludge/thrombus at admission (*P*=0.031 and *P*<0.0001 respectively). Receiver operating characteristic curve analysis revealed the optimal cut-off for 3D ES LAAV to discriminate patients at risk of death within 2 years was 9.3 mL. Kaplan–Meier curves demonstrated a significant difference in survival at 2-year follow-up according to this value: 3 deaths occurred in the group with 3D ES LAAV <9.3mL and 11 in those with volume ≥9.3 mL (*P*=0.02).

**CONCLUSIONS:** 3D characterization of LAAV depicts a degree of LAA remodeling in AF that appears associated with LA/LAA thrombogenicity and mid-term prognosis.

**CONDENSED ABSTRACT:** Limited data are available on left atrial appendage (LAA) remodeling in atrial fibrillation (AF). We hypothesized that 3-dimensional (3D) evaluation of the LAA volume in AF could help to refine the prothrombotic state and prognosis in AF. Patients with LAA sludge and/or thrombus exhibited lower LAA filling and emptying flow velocities, and higher 2-dimensional (2D) and 3D LAA measurements. On multivariable analysis, LAA filling flow velocity and 3D end-systolic LAA volume were associated with the presence of LAA sludge/thrombus at admission (respectively, *P*=0.031 and *P*<0.0001). Kaplan–Meier curves demonstrated a significant difference in survival at 2 years according to 3D ES LAA volume (*P*=0.02). Three dimensional LAA volume reflects the degree of LAA remodeling in AF and is associated with prothrombotic state and prognosis.

## INTRODUCTION

A prothrombotic state in atrial fibrillation (AF) fulfils Virchow’s triad of blood hypercoagulability, endothelial injury and abnormal blood stasis.^1^ Left atrial (LA) blood stasis (i.e., transesophageal echocardiography [TEE]-detected reduced LA appendage [LAA] blood flow, LAA spontaneous echo contrast, sludge) occurs as a direct consequence of AF and contributes to thrombus formation. The thromboembolism risk assessment is essential in the management of AF.^2^

Cardiac imaging plays a central role in evaluating the AF substrate (i.e., atrial cardiomyopathy).^3, 4^ Two-dimensional (2D)/3-dimensional (3D) transthoracic echocardiography (TTE), TEE, cardiovascular magnetic resonance imaging and computed tomography are useful imaging modalities for the volumetric and functional assessment of atrial remodeling.^5^ Several cardiac imaging parameters have been described as markers of cardioembolic risk: large LA volumes,^6^ left ventricular systolic dysfunction,^7^ TEE-detected LA spontaneous echo contrast,^8^ decreased LAA emptying and filling velocities,^9^ and presence of aortic atheroma.^10^

Limited data are available on the association between LAA parameters and LAA prothrombotic state in computed tomography, magnetic resonance imaging^11–13^ and TEE ^14–16^ (Table S1). While LA volume appears associated with patient prognosis,^17^ little is known about the relationship between LAA parameters and LAA prothrombotic state and prognosis.

The aims of this study in an AF population were threefold: 1) to provide a comprehensive evaluation of LAA using 2D and 3D TEE; 2) to determine which LAA parameter is best associated with a prothrombotic state; and 3) to study the relationship between LAA-derived parameters and prognosis.

## METHODS

### Study Design and Population

FASTRHAC is a multicenter, prospective study of patients hospitalized for AF (Ethics committee authorization: CPP Ile de France V, number: 2014-A00280-47. NCT 02741349). The FASTRHAC methods have been described previously.^5, 18^ All consecutive patients (≥18 years) hospitalized for paroxysmal or persistent AF diagnosed on a 12-lead electrocardiogram who provided written informed consent were included. Exclusion criteria were organic valvular disease defined according to the guidelines,^19–21^ presence of a mechanical or biological prosthesis, contraindication to anticoagulant treatment, lack of affiliation to a social security regimen, severe psychiatric history, and subjects considered unlikely to present for follow-up. Comprehensive clinical characteristics, biological variables, and 2-D and 3D TTE and TEE data were collected over 2 years. CHA_2_DS_2_-VASc score (Congestive heart failure, Hypertension, Age ≥75 [doubled], Diabetes mellitus, prior Stroke or transient ischemic attack or thromboembolism [doubled], Vascular disease, Age 65 to 74, Sex category [female]) was determined in each patient.

This analysis focuses on the first 218 consecutive patients enrolled in the study who underwent 3D TEE at admission and had a comprehensive follow-up of 2 years (Figure 1).

**Figure 1.**
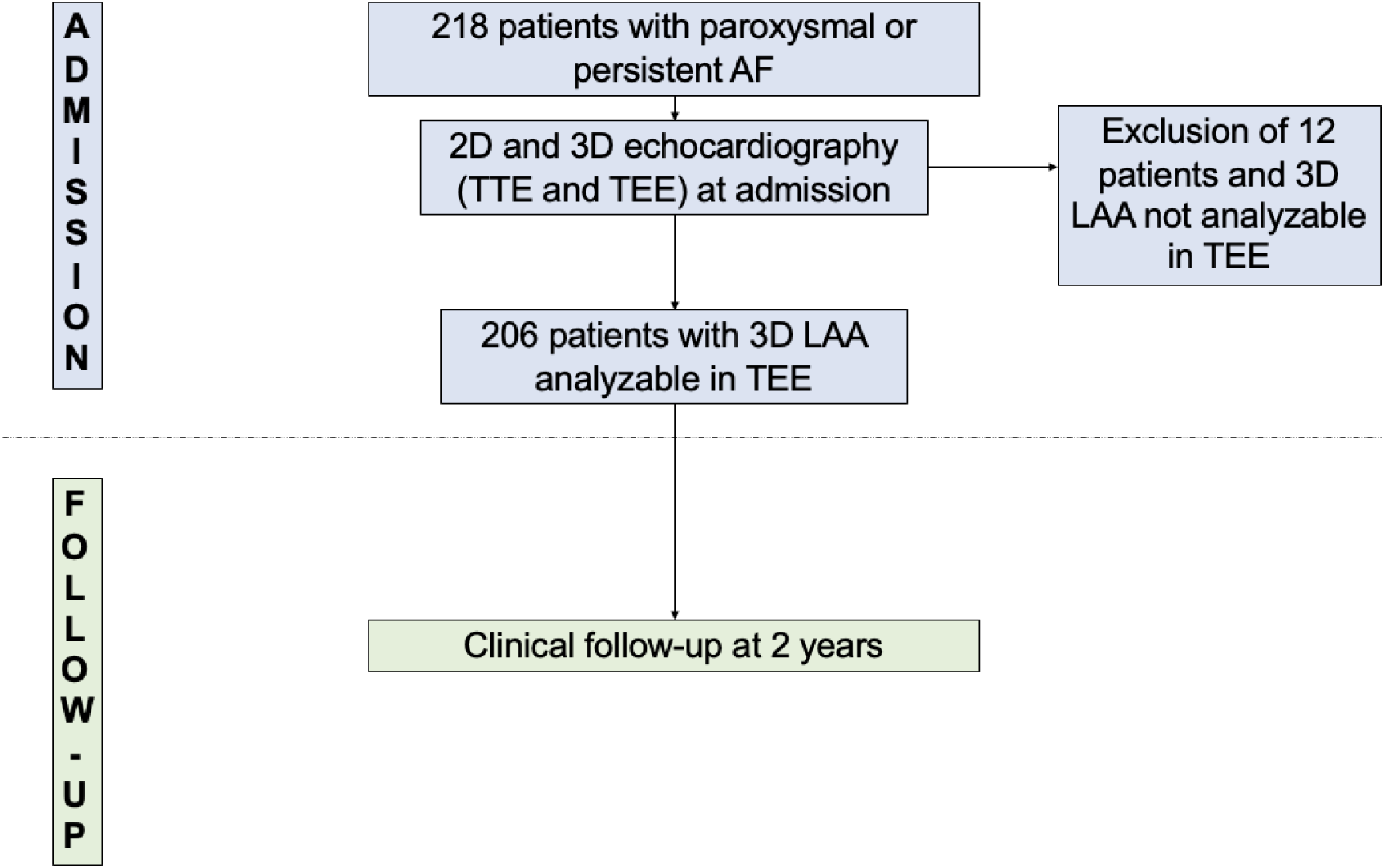
Study flow chart. 3D indicates 3-dimensional; AF, atrial fibrillation; LAA, left atrial appendage; TEE, transesophageal echocardiography; and TTE, transthoracic echocardiography.

### Transthoracic and Transesophageal Echocardiography

2D and 3D TTE and TEE were performed within 24 hours of admission by experienced cardiologists using X5-1, X72T and X82T transducers connected to an EPIQ 7 or CVx (Philips Medical Systems, Andover, MA, USA). The data were transferred and analyzed offline using a TOMTEC workstation (Image Arena; TOMTEC, Unterschleissheim, Germany) (L.S.D., T.S., E.C. and C.A.). 2D and 3D measurements were analyzed following US and European Chamber Quantitation Guidelines.^22, 23^ Volumetric measurements were indexed to body surface area.

2D transesophageal evaluation of the LAA included the following parameters (Figure 2): evaluation of spontaneous echo contrast between 0° and 120° according to the Fatkin classification^8^ (grades 0, 1, 2, 3, sludge, thrombus); measurements at 90° of 2D ES and ED LAA ostium diameter, 2D ES and ED LAA area; measurements at 90° of LAA emptying and filling flow velocities; and evaluation of trabeculation severity (mild, moderate, severe).

**Figure 2.**
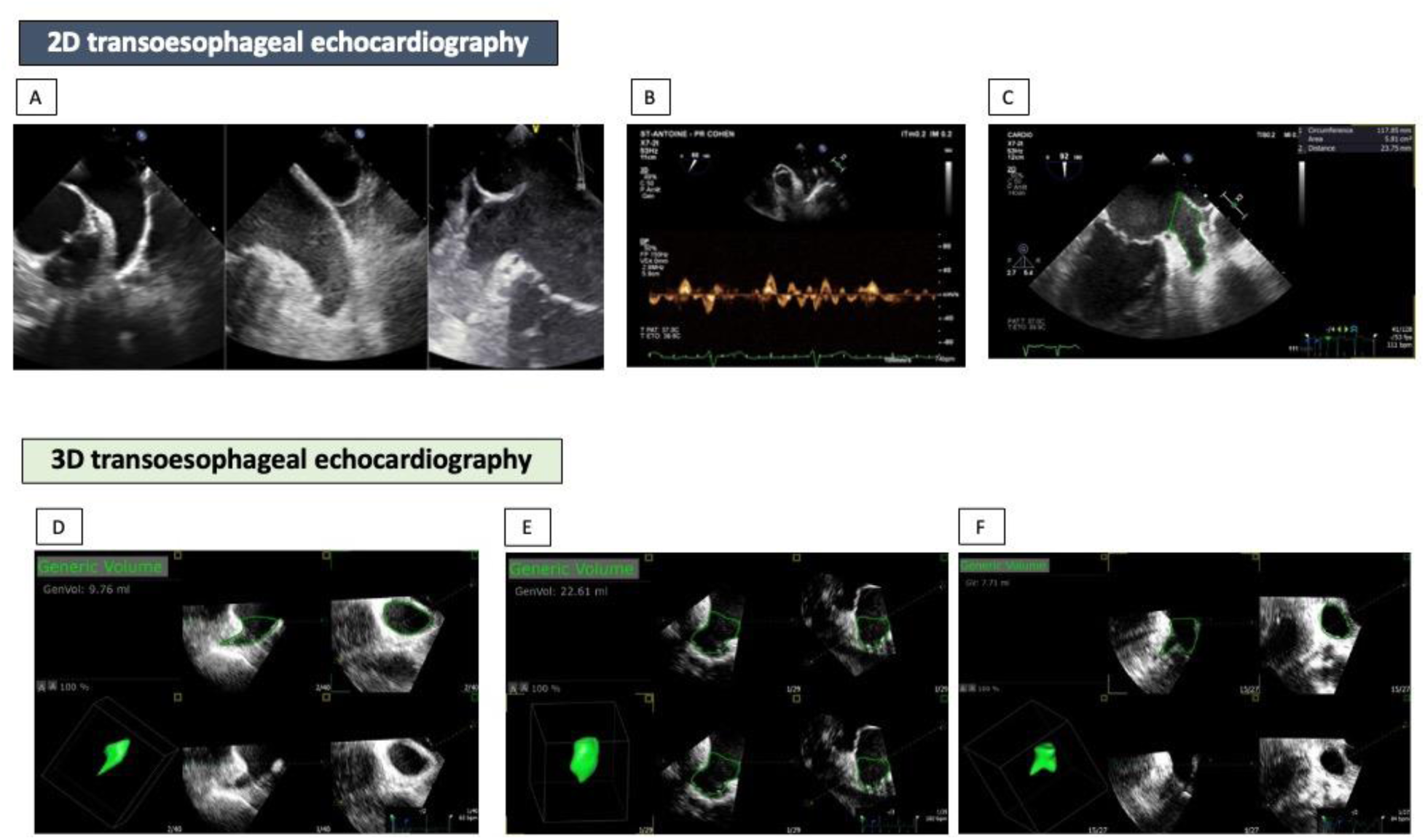
Evaluation of LAA in 2D (A, B, C) and 3D (D, E, F) TEE. A: examples of LA/LAA spontaneous echo contrast grade 0 (left), sludge (middle), thrombus (right) according to Fatkin classification^8^; B: evaluation of LAA flow velocities; C: 2D measurements of the LAA area at 92° (end-systolic left atrial appendage area and ostium diameter). Examples of different LAA morphologies evaluated with Zoom 3D mode 1 beat; D: left atrial appendage with chicken-wing morphology and a volume of 9.76 mL; E: left atrial appendage with no chicken-wing morphology and a volume of 22.61 mL; and F: left atrial appendage with 2 lobes and a volume of 7.71 mL. 2D indicates 2-dimensional; 3D, 3-dimensional; AF, atrial fibrillation; LA, left atrial; LAA, left atrial appendage; and TEE, transesophageal echocardiography.

The analysis of LAA with 3D TEE used 3D 1-beat zoom mode. 3D datasets of the LAA were deemed adequate for analysis if all cavity segments were visible in the dynamic dataset. 3D full-volume datasets were analyzed using software specifically designed for 3D volumetric analysis (Tomtec, Generic Volume Software). The 3D TEE evaluation of the LAA included the following parameters (Figure 2): ES and ED LAA volumes (LAAV); ES and ED LAA ostium areas; LAA morphology; and LAA number of lobes.

### Statistical Analysis

The study population was divided into 2 groups according to the presence or absence of sludge and/or thrombus in the LAA. Then, using the non-parametric Kruskal−Wallis rank test for continuous variables and the χ^2^ or Fisher’s exact test for categorical variables, the baseline parameters were compared between the 2 groups. First, logistic regression analyses were performed to identify determinants of the presence of sludge and/or thrombus in the LAA. For each continuous variable, the choice between a continuous or categorical classification was based on the lowest value of Akaike’s information criterion for the corresponding univariate Cox model. To avoid collinearity and overfitting problems, each clinical and biological variable was entered into a stepwise backward multivariable model and variables with *P*<0.20 were retained. The same method was repeated for the echocardiographic variables (Table S2). Finally, a multivariable model adjusted for the selected variables was constructed on the population of interest.

Second, a receiver operating characteristic curve was depicted to analyze the capacity of the 3D ES LAAV to identify patients at risk of dying within 2 years. A cut-off of 9.3 mL was determined (Figure S1) and was used to build the Kaplan−Meir survival curves and was compared using the log-rank test. Cox regression models analyses were performed to identify variables associated with death within 24 months of follow-up.

Biological characterization of the population with a C-reactive protein (CRP) value >10 mg/L and B-type natriuretic peptide (BNP) value ≥400 pg/mL were based on a careful analysis of the literature.^24, 25^

All analyses were performed using STATA V12 (StataCorp, College Station, TX). *P*<0.05 was considered statistically significant.

## Results

### Patient Baseline Characteristics

3D LAA evaluation was feasible in 206 of 218 patients undergoing TEE (94.5%) (Figure 1). At admission, LAA spontaneous echo contrast grade 0 was found in 85 (41.3%) patients, grade 1 in 47 (22.8%), grade 2 in 26 (12.6%), grade 3 in 15 (7.3%), sludge in 33 (16.0%) and thrombus in 7 (3.4%). One-hundred and twenty-nine (62.6%) patients were male, and the median age was 66.3±11.5 years; 75.7% patients had a CHA_2_DS_2_-VASc ≥2, 30.1% had paroxysmal AF and 69.9% had persistent AF at admission (Table 1).

**Table 1.** Clinical and Biological Characteristics in the Global Population and According to the presence of LAA Sludge and/or Thrombus at admission.

| Variable | All<br>(n=206) | No LAA sludge<br>and/or thrombus<br>(n=171) | LAA sludge and/or<br>thrombus<br>(n=35) | *P* |
| --- | --- | --- | --- | --- |
| Clinical characteristics and cardiovascular risk factors |  |  |  |  |
| Sex, male | 129 (62.6) | 105 (61.4) | 24 (68.6) | 0.43 |
| Age, years | 66.3±11.5 | 66.3±11.7 | 66.1±10.1 | 0.88 |
| Body mass index, kg/m<sup>2</sup> | 27.0 (23.6–30.9) | 26.9 (23.2–31.0) | 27.1 (23.9–29.5) | 0.66 |
| Hypertension | 110 (53.4) | 92 (53.8) | 18 (51.4) | 0.80 |
| Diabetes | 39 (18.9) | 34 (19.9) | 5 (14.3) | 0.44 |
| Hypercholesterolemia | 69 (33.5) | 57 (33.3) | 12 (34.3) | 0.91 |
| Current smoker | 37 (18.0) | 32 (18.7) | 5 (14.3) | 0.53 |
| Medical history |  |  |  |  |
| AF | 72 (35.0) | 58 (33.9) | 14 (40.0) | 0.49 |
| Heart failure | 32 (15.5) | 21 (12.3) | 11 (31.4) | 0.004 |
| Stroke | 18 (8.7) | 14 (8.2) | 4 (11.4) | 0.54 |
| Myocardial infarction | 30 (14.6) | 23 (13.5) | 7 (20.0) | 0.32 |
| Renal failure | 17 (8.3) | 15 (8.8) | 2 (5.7) | 0.55 |
| Reason for hospitalization |  |  |  |  |
| AF without heart failure | 133 (64.6) | 117 (68.4) | 16 (45.7) | 0.011 |
| AF with heart failure | 73 (35.4) | 54 (31.6) | 19 (54.3) |  |
| AF classification |  |  |  |  |
| Paroxysmal | 62 (30.1) | 58 (33.9) | 4 (11.4) | 0.008 |
| Persistent | 144 (69.9) | 113 (66.1) | 31 (88.6) |  |
| CHA<sub>2</sub>DS<sub>2</sub>-VASc score |  |  |  |  |
| 0 | 15 (7.3) | 13 (7.6) | 2 (5.7) | 0.12 |
| 1 | 35 (17.0) | 33 (19.3) | 2 (5.7) |  |
| ≥2 | 156 (75.7) | 125 (73.1) | 31 (88.6) |  |
| Biology |  |  |  |  |
| B-type natriuretic peptide,<br>pg/mL | n=205<br>238 (123–378) | n=170<br>203 (104–317) | 446 (171–759) | 0.0001 |
| Troponin, ng/mL | n=205<br>0.04 (0.04–0.04) | n=170<br>0.04 (0.04–0.04) | 0.04 (0.4–0.04) | 0.77 |
| D dimers, ng/mL | n=195<br>337 (270–543) | n=162<br>325 (270–551) | n=33<br>424 (283–690) | 0.18 |
| Glomerular filtration rate*,<br>mL/min/1.73 m<sup>2</sup> | 75.7 (63.4–89.2) | 75.5 (63.1–90.2) | 76.0 (65.1–87.3) | 0.99 |
| C-reactive protein, mg/L | 4.0 (3.0–10.0) | 3.3 (3.0–7.2) | 6.2 (3.0–15.0) | 0.025 |
| Low-density lipoprotein<br>cholesterol, g/L | n=204<br>1.04 (0.78–1.23) | n=169<br>1.03 (0.78–1.20) | 1.08 (0.76–1.32) | 0.52 |
| Glycated hemoglobin A<sub>1C</sub>,<br>% | n=194<br>5.8 (5.5–6.2) | n=160<br>5.7 (5.5–6.1) | n=34<br>6.1 (5.6–6.6) | 0.08 |

Values presented as count (%) or median (interquartile range).
\*Calculated with the Modification of Diet in Renal Disease equation.
AF indicates atrial fibrillation; and LAA, left atrial appendage.

Thirty-five (17.0%) patients had LAA sludge/thrombus at admission (Table 1). Patients with LAA sludge/thrombus had a higher prevalence of heart failure (and hospitalization due to AF associated with heart failure). Patients with sludge and/or thrombus had higher values for BNP and CRP. There were no significant differences between the groups regarding clinical and cardiovascular risk factors, CHA_2_DS_2_-VASc score, or troponin and D-dimer values.

Echocardiographic characteristics are shown in Table 2. On 2D TTE, patients with LAA sludge/thrombus had higher median LA volume and lower median LVEF than those without. On 2D TEE, patients with LAA sludge/thrombus had lower LAA emptying and filling flow velocities and greater LAA measurements (ostium diameter, area). On 3D TEE, patients with LAA sludge/thrombus had a higher median LAAV, greater ED LAA ostium area, and more frequently exhibited a non-chicken wing morphology.

**Table 2.** Echocardiography Characteristics in the Global Population and According to the Presence or Absence of LAA Sludge and/or Thrombus at Admission.

| Variable | All<br>(n=206) | No LAA sludge<br>and/or thrombus<br>(n=171) | LAA sludge and/or<br>thrombus<br>(n=35) | P |
| --- | --- | --- | --- | --- |
| 2D TTE |  |  |  |  |
| LVEF*, % | 52.00 (38.4–60.0) | 54.9 (44.8–60.0) | 37.0 (21.0–50.0) | 0.0001 |
| ES LAV, mL (n=204) | 86.1 (69.2–104.5) | 80.8 (68.0–99.8) | 99.4 (83.6–115.0) | 0.0005 |
| ES LAVi, mL/m<sup>2</sup> (n=204) | 45.0 (36.2–53.9) | 42.8 (35.3–51.4) | 51.5 (45.1–56.4) | 0.0004 |
| PAP, mmHg (n=168) | 29 (24–36) | 29 (24–35) | 28 (24–36) | 0.97 |
| 2D TEE |  |  |  |  |
| LAA emptying flow velocity, cm/s<br>(n=199) | 32 (23–49) | 35 (26–52) | 21 (17–28) | 0.0001 |
| LAA filling flow velocity, cm/s (n=199) | 35 (24–50) | 38 (29–53) | 22 (19–31) | 0.0001 |
| ES LAA ostium diameter, mm | 20.82 (17.78–23.65) | 20.2 (17.5–23.6) | 23.1 (20.9–25.5) | 0.004 |
| ES LAA area, cm<sup>2</sup> | 5.98 (4.46–7.94) | 5.9 (4.2–7.7) | 7.2 (5.4–9.2) | 0.02 |
| ED LAA ostium diameter, mm | 18.70 (15.10–22.50) | 17.7 (14.7–22.0) | 21.6 (18.3–24.4) | 0.002 |
| ED LAA area, cm<sup>2</sup> | 4.95 (3.57–7.22) | 4.7 (3.4–7.1) | 6.4 (4.6–8.7) | 0.002 |
| Trabeculations (n=203) |  |  |  | 0.16 |
| Mild | 102 (50.3) | 89 (53.0) | 13 (37.1) |  |
| Moderate | 81 (39.9) | 62 (36.9) | 19 (54.3) |  |
| Severe | 20 (9.9) | 17 (10.1) | 3 (8.6) |  |
| Aortic atheroma (n=205) | 80 (39.0) | 67 (39.2) | 13 (38.2) | 0.92 |
| 3D TEE |  |  |  |  |
| ES LAAV, mL | 9.02 (6.96–12.15) | 8.7 (6.8–11.9) | 10.9 (8.7–14.8) | 0.003 |
| ES LAA ostium area, mm<sup>2</sup> (n=205) | 3.90 (3.13–4.98) | 3.9 (3.1–4.9) | 4.5 (3.2–6.1) | 0.05 |
| ED LAAV, mL | 7.93 (5.98–10.87) | 7.8 (5.6–10.6) | 10.0 (7.6–13.0) | 0.002 |

| Variable | All<br>(n=206) | No LAA sludge<br>and/or thrombus<br>(n=171) | LAA sludge and/or<br>thrombus<br>(n=35) | *P* |
| --- | --- | --- | --- | --- |
| ED LAA ostium area, mm<sup>2</sup> (n=205) | 3.48 (2.69–4.53) | 3.4 (2.6–4.3) | 4.1 (3.1–6.0) | 0.003 |
| LAA morphology (n=205) |  |  |  | 0.007 |
| Chicken wing | 122 (59.5) | 109 (63.7) | 13 (38.2) |  |
| Non-chicken wing | 83 (40.5) | 62 (36.3) | 21 (61.8) |  |
| Number of lobes (n=205) |  |  |  | 0.56 |
| 1 | 94 (45.9) | 79 (46.2) | 15 (44.1) |  |
| 2 | 78 (38.1) | 62 (36.3) | 16 (47.1) |  |
| 3 | 25 (12.2) | 23 (13.5) | 2 (5.9) |  |
| 4 | 8 (3.9) | 7 (4.1) | 1 (2.9) |  |

Values presented as count (%) or median (interquartile range).
\*Calculated using the Simpson method.
AF indicates atrial fibrillation; ED, end diastolic; ES, end systolic; LAA, left atrial appendage;
LAAV, left atrial appendage volume; LVEF, left ventricular ejection fraction; TEE, transesophageal
echocardiography; and TTE, transthoracic echocardiography.

### Determinants of LAA Prothrombotic State

On univariable analysis, predictors of LAA sludge/thrombus at admission were: history of heart failure or acute heart failure, persistent AF, hospitalization for AF with heart failure, CRP >10 mg/L, BNP ≥400 pg/mL, decreased LVEF, LAVI ≥45 mL/m^2^, decreased LAA filling flow velocity, greater 2D LAA measurements (LAA ostium diameter, LAA area), increased 3D LAA measurements (LAAV, ostium area) and non-chicken wing morphology (Table S3). On multivariable analysis, LAA filling flow velocity and 3D ES LAAV were associated with LAA sludge/thrombus at admission (Table 3).

**Table 3.**
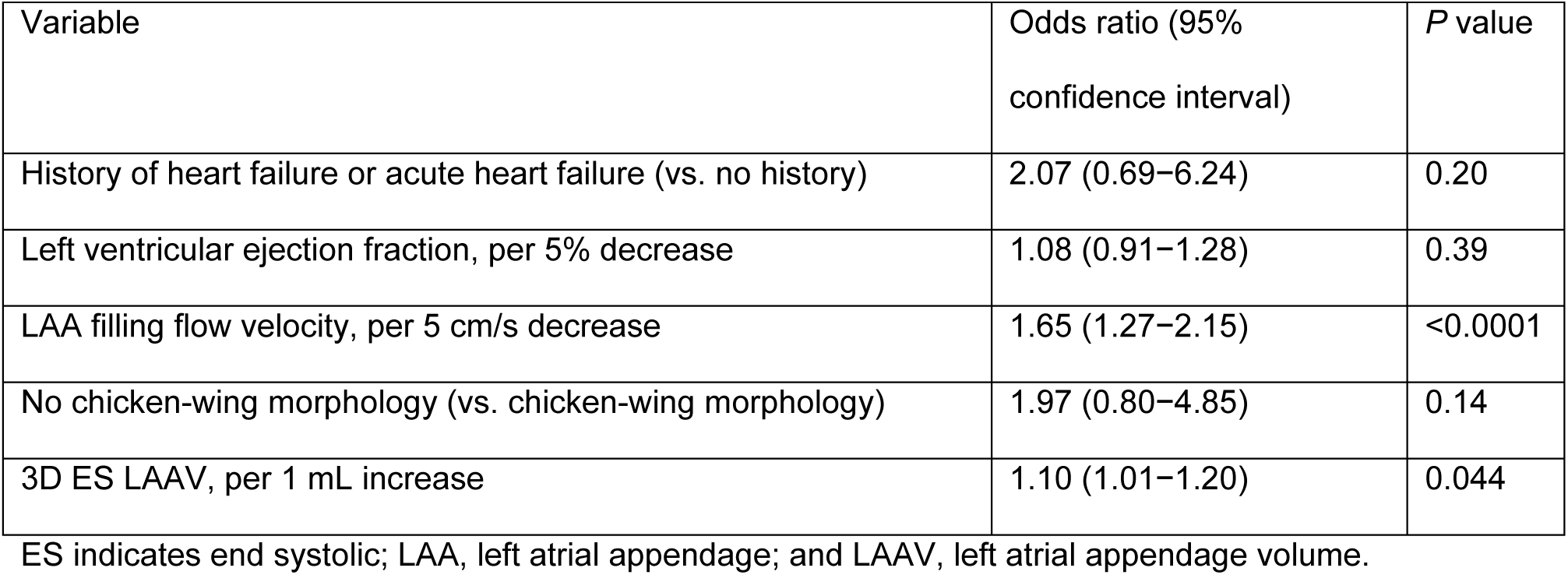
Determinants of Prothrombotic State on Multivariable Logistic Regression Analysis (n=198)

| Variable | Odds ratio (95% confidence interval) | *P* value |
| --- | --- | --- |
| History of heart failure or acute heart failure (vs. no history) | 2.07 (0.69–6.24) | 0.20 |
| Left ventricular ejection fraction, per 5% decrease | 1.08 (0.91–1.28) | 0.39 |
| LAA filling flow velocity, per 5 cm/s decrease | 1.65 (1.27–2.15) | <0.0001 |
| No chicken-wing morphology (vs. chicken-wing morphology) | 1.97 (0.80–4.85) | 0.14 |
| 3D ES LAAV, per 1 mL increase | 1.10 (1.01–1.20) | 0.044 |

ES indicates end systolic; LAA, left atrial appendage; and LAAV, left atrial appendage volume.

### Clinical, Biological and Echocardiography Characteristics According to 3D ES LAAV

Receiver operating characteristic curve analysis revealed that the optimal cut-off for 3D ES LAAV to discriminate patients at risk of death within 2 years was 9.3 mL (area under the curve 0.61±0.05; 95% confidence interval 0.50–0.72) (Figure S1).

Patients with LAAV ≥9.3 mL were more frequently male, had a history of heart failure or renal failure and were hospitalized for AF associated with heart failure. They had also higher values for BNP, CRP and glycated hemoglobin A_1c_ (Table S4). Patients with LAAV ≥9.3 mL had a lower median LVEF, greater LA volumes, more severe grade of LAA sludge and/or thrombus, lower LAA emptying flow velocity, and greater 2D LAA measurements (ostium diameter, area). In 3D TEE, patients with LAAV ≥9.3 mL had greater 3D LAA measurements (volume, ostium area) and no significant difference in LAA morphology including number of lobes (Table S5).

### Clinical, Biological and Echocardiography Characteristics According to AF Pattern

Patients with persistent AF had a higher body mass index, higher prevalence’s of hypertension and diabetes, history of AF, heart failure, renal failure, hospitalization for AF and heart failure, a higher CHA_2_DS_2_-VASc score, higher BNP, CRP, and HbA_1c_ values, and lower glomerular filtration rate (Table S6).

Echocardiographic analysis showed that patients with persistent AF had a lower LVEF, higher LAV and pulmonary artery pressures, higher degree of LAA spontaneous echo contrast, higher rates of LAA sludge and/or thrombus, lower LAA emptying and filling flow velocities, greater 2D LAA area and ostium diameter, and greater 3D LAAV and ostium area (Table S7).

### Clinical Events During 2 Years of Follow-Up

Over a mean ± standard deviation follow-up of 22.0±5.4 months, 56 (27.2%) patients had recurrent AF, 23 (11.2%) had heart failure, 5 (2.4%) myocardial infarction, 2 (1.0%) stroke, and 14 (6.8%) died (11 cardiovascular deaths). Kaplan–Meier curves demonstrated a significant difference in survival at 2 years according to 3D ES LAAV: 3 deaths occurred in the group with 3D ES LAAV <9.3 mL and 11 in the group with a volume ≥9.3 mL (*P*=0.02) (Figure 3).

**Figure 3.**
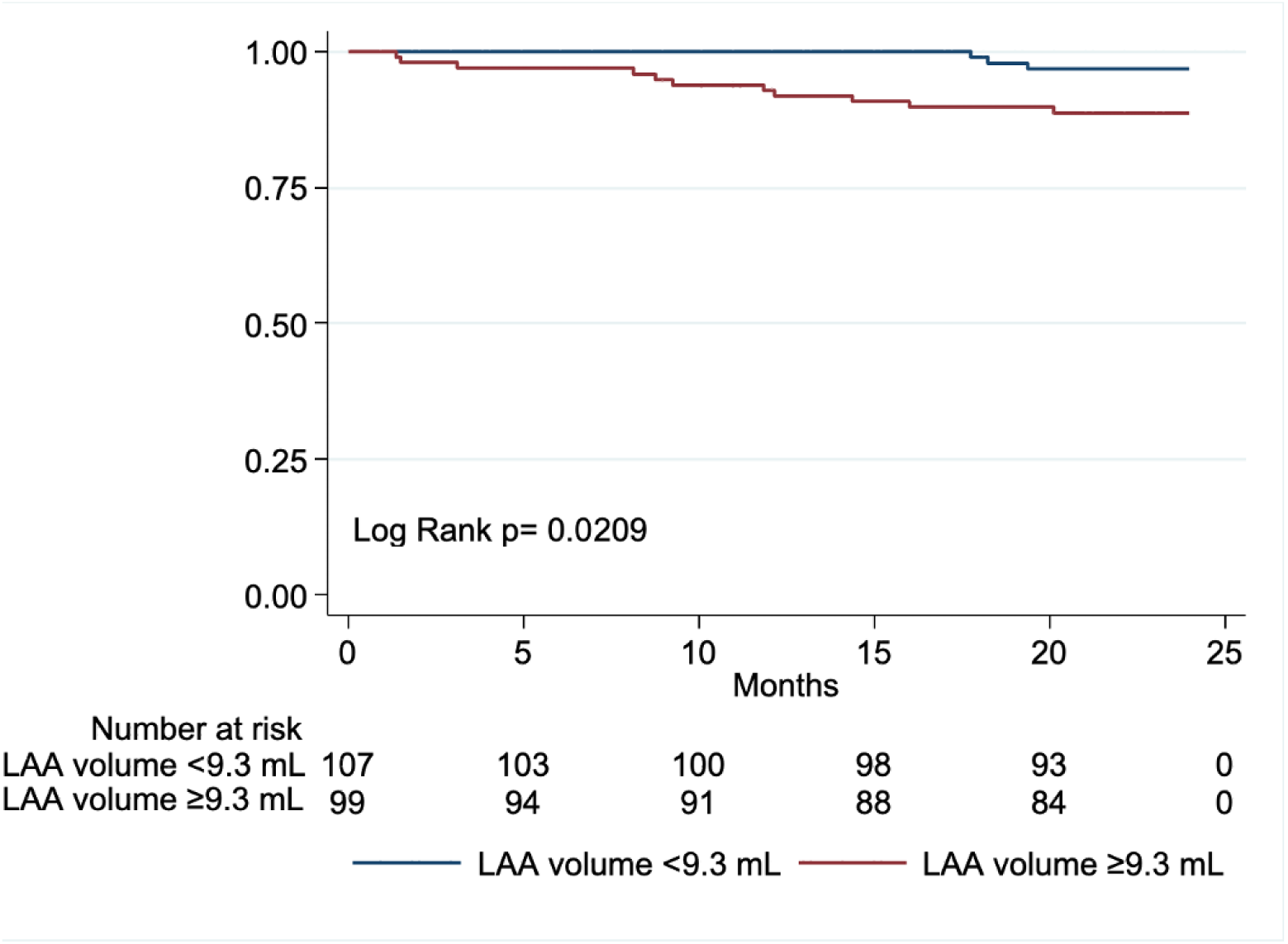
Kaplan–Meier Survival Estimates According to 3D ES LAA Volume at 2 Years. Kaplan–Meier curves showing event-free survival according to the 3D ES LAA Volume <9.3 mL (blue) or ≥9.3 mL (red). 3D indicates 3-dimensional; ES, end-systolic; and LAA, left atrial appendage.

On univariable analysis, female sex, history of heart failure, presence of heart failure at admission, CRP >10 mg/L, low LVEF, high left atrial volume index, grade 2 spontaneous echo contrast, low LAA filling and emptying velocities, and LAAV ≥9.3 mL were predictors of death (Table S8). On multivariable analysis, CRP >10 mg/L was associated with death (Table 4).

**Table 4.** Multivariable Cox Model for Prediction of Death at 2-year Follow-Up (n=199)

| Variable | Multivariable Cox model<br>HR (95% CI) | *P* |
| --- | --- | --- |
| Female sex (vs. male sex) | 0.15 (0.02–1.17) | 0.07 |
| AF with heart failure (vs. no heart failure) | 4.79 (0.86–26.72) | 0.07 |
| Renal failure (vs. no renal failure) | 3.53 (0.64–19.38) | 0.15 |
| CRP >10 mg/L (vs. <10 mg/L) | 3.43 (1.06–11.16) | 0.04 |
| LVEF, per 5 % decrease | 1.16 (0.93–1.44) | 0.18 |
| LAA emptying flow velocity <25 cm/s (vs. ≥25 cm/s) | 2.74 (0.86–8.72) | 0.09 |
| 3D ES LAAV ≥9.3 mL (vs. <9.3 mL) | 1.45 (0.38–5.59) | 0.59 |

AF indicates atrial fibrillation; CI, confidence interval; CRP, C-reactive protein; ES, end systolic; HR, hazard ratio; LAA, left atrial appendage; LAAV, left atrial appendage volume; and LVEF, left ventricular ejection fraction.

## DISCUSSION

The findings from our study suggest that increased 3D LAAV is associated with LAA prothrombotic markers and risk of death. On multivariable analysis, LAA filling flow velocity and 3D ES LAAV were the only determinants associated with LAA sludge/thrombus, independently of clinical, biological, and other echocardiographic parameters. Kaplan–Meier curves demonstrated a significant difference in survival at 2 years when using 3D ES LAAV.

### 3D LAAV: A New Parameter to Evaluate Prothrombotic State in AF

Several parameters have been described using echocardiography to evaluate LAA prothrombotic state in AF. In TEE, LA/LAA spontaneous echo contrast^8^ and LAA emptying or filling velocities^9^ are the most commonly validated and used parameters. In the literature, 3D LAA morphology (chicken wing, windsock, cactus, cauliflower) is also associated with thromboembolic history, with a higher thromboembolic risk for non−chicken-wing morphology.^13^ However, LAA morphology is a subjective evaluation. In our study, 3D ES LAAV was more accurately associated with thromboembolic state (on multivariable analysis) (Table 3) when compared with 3D LAA morphology (on univariable analysis) (Table S3). Thus, we propose to consider 3D LAAV as an additional pertinent parameter to evaluate the LAA thrombogenic state in AF.

### LAA Remodeling in AF

In comparison with healthy individuals, patients with AF had higher LAAV.^14^ The increase in LAAV may correlate with the degree with LAA dysfunction, and be an integral part of the atrial cardiomyopathy concept, defined as “structural, architectural, contractile or electrophysiological changes affecting the atria with the potential to produce clinically-relevant manifestations”.^4^ In a recent study, LAA fibrosis and endothelial damage were associated with LAA thrombus and stroke, independently of clinical factors.^26^ It is interesting to note the association between 3D LAAV and clinical patterns of AF; in our study, patients with paroxysmal AF had a lower median LAAV compared to those with persistent AF (Table S7). These results are consistent with the literature: patients with paroxysmal AF had fewer thromboembolic events and deaths compared to those with persistent or permanent AF.^27^

Recently reported data indicate that early rhythm control reduces cardiovascular death, stroke, hospitalization for heart failure and acute coronary syndrome across AF clinical patterns.^28^ We demonstrated that management of AF focused on restoration of sinus rhythm could induce anatomical and/or functional cardiac cavity reverse remodeling.^5^ The integration of LAA remodeling could be a key future step in understanding the pathophysiological processes involved in AF.

### Clinical events

Over 2 years of follow-up, 14 deaths (11 cardiovascular) and 2 strokes occurred, highlighting the effectiveness of AF management in the prevention of strokes. LAAV is associated with the prothrombotic state (LAA sludge/thrombus) independently of clinical and biological factors. Furthermore, in Kaplan−Meier analysis, 3D LAAV was associated with death in the univariable analysis. Thus, LAAV appears to be a key parameter of atrial remodeling and could be integrated into the evaluation of thromboembolic risk (Figure 4).

**Figure 4.**
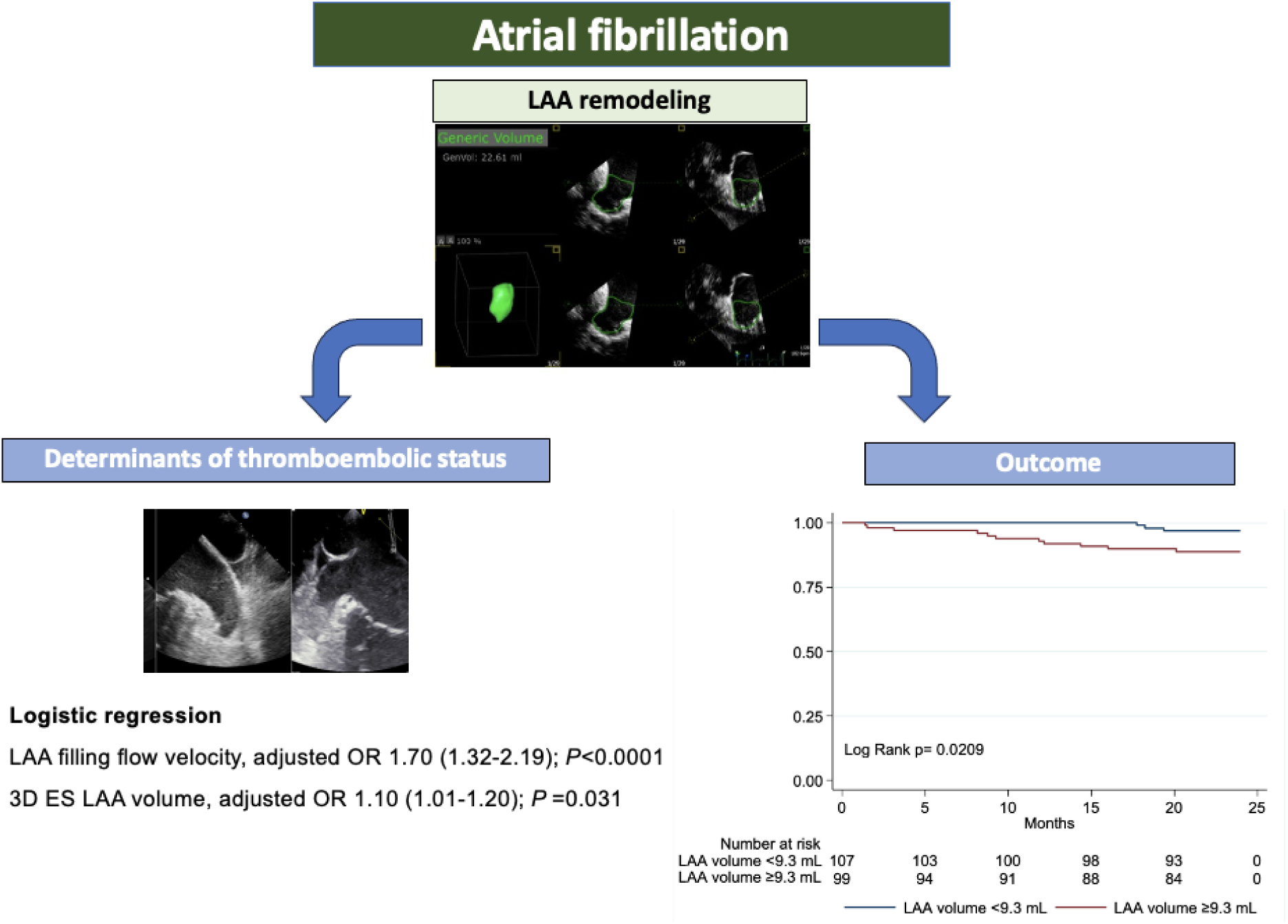
Role of 3D of LAA in AF: Association with Prothrombotic State and Prognosis. 3D indicates 3-dimensional; ES, end-systolic; LAA, left atrial appendage; and OR, odds ratio.

### Limitations

In cardiac imaging, the irregular rhythm in AF always presents a challenge for image acquisition and quantitation. However, accurate 3D evaluation of LAAV was possible in 206 (94.0%) examinations. It would have been interesting to use an additional imaging technique to measure LAAV (e.g., computed tomography or cardiac magnetic resonance imaging); however, our study was performed as part of routine care and did not benefit from examinations not indicated in the patient’s usual management. In this study, we described LAA morphology in 2 stages (chicken wing versus non-chicken wing). In our practice, this 2-stage evaluation appears to reduce the subjectivity of LAA morphology evaluation in comparison with the multiple morphology assessment (chicken wing, cauliflower, cactus, windsock). In addition to the LAA morphology evaluation, we suggest that LAAV evaluation is a more objective parameter. In our study, the reason for hospitalization was heart failure and AF in 35.4%, and we cannot exclude that the LAAV could have been modified by changes in filling pressure in the heart failure and AF group. Finally, our population included patients with paroxysmal or persistent AF; the absence of permanent AF does not allow us to extend our results to this population.

## Conclusions

3D evaluation of LAAV is a new tool for evaluation of thromboembolic state and prognosis in patients with AF, which appears to be more accurate than LAA morphology for the evaluation of prothrombotic state. LAAV could be integrated into the characterization of LA/LAA remodeling during AF, and into the concept of atrial cardiomyopathy as a pertinent diagnostic tool. Further investigations are necessary to understand the role of LAA remodeling in the thrombogenic function of LAA as well as in the long-term prognosis in patients with AF.

## Data Availability

All data referred to the mauscript are available

## Acknowledgments

Sophie Rushton-Smith, PhD (Medlink Healthcare Communications, London), provided editorial assistance including editing, checking content and language and formatting, and was funded by the authors.

## Supplemental Material

Tables S1–S8

Figure S1

References 1–6

## Clinical Perspective

### What Is New?

Three-dimensional evaluation of left atrial appendage volume is a useful new tool for evaluating the thromboembolic state and prognosis in patients in atrial fibrillation.

### What Are the Clinical Implications?

Further investigations are needed to clarify the integration of left atrial appendage volume into the imaging diagnostic criteria in atrial cardiomyopathy.

## Non-standard Abbreviations and Acronyms

2D: 2-dimensional
3D: 3-dimensional
AF: atrial fibrillation
BNP: B-type natriuretic peptide
CRP: C-reactive protein
ED: end-diastole
ES: end-systole
I: indexed
LAA: left atrial appendage
LAAV: left atrial appendage volume
TEE: transesophageal echocardiography
TTE: transthoracic echocardiography

